# Prediction of future chronic hypertension from maternal characteristics in early pregnancy

**DOI:** 10.1101/2023.04.26.23289181

**Authors:** Marietta Charakida, Alan Wright, Laura A Magee, Argyro Syngelaki, Peter von Dadelszen, Ranjit Akolekar, David Wright, Kypros H Nicolaides

## Abstract

**Background:** Pre-eclampsia (PE) and gestational hypertension (GH) identify women at increased risk of chronic hypertension (CH) and cardiovascular disease, but as efforts to prevent PE and GH advance, fewer women at increased cardiovascular risk will be identified.

**Methods:** Cohort of 26,511 women seen in two consecutive pregnancies. Included were women without CH, with information on maternal characteristics and blood pressure (BP) at 11-13 weeks’ gestation, and development of PE or GH in the index pregnancy. Logistic regression models were fitted for prediction of development of future CH by the 20^th^ week of the subsequent pregnancy. Performance of screening and risk calibration of the model were assessed.

**Results:** 1560 (5.9%) women developed PE or GH (index pregnancy), and 215 (0.8%) developed future CH, a median of 3.0 years later. Predictors from the index pregnancy of development of future CH were: early pregnancy maternal age, weight and BP; Black or South Asian ethnicity; family history of PE; parity; and development of PE or GH. PE or GH accounted for 52.1% (95% confidence interval 45.2-58.9%) of future CH. At a screen-positive-rate of 10%, a model including terms for maternal characteristics and early pregnancy BP accounted for 67.9% (61.2-74.5) of future CH; addition of the development of PE or GH detected 73.5% (67.1-79.3) of future CH. Risks produced from the predictive model were well-calibrated and confirmed by five-fold cross-validation.

**Conclusion:** Early maternal characteristics and BP are effective in predicting development of future CH. As new interventions are expected to reduce the occurrence of PE and GH, our study results offer an alternative strategy for identifying women at increased risk of future CH and are applicable worldwide.

## INTRODUCTION

Cardiovascular (CV) disease is the leading cause of mortality among women, exceeding even breast cancer.^1^ Hypertension is the leading modifiable CV risk factor that is rising in incidence, globally;^2^ a 10mm Hg increase in systolic blood pressure (BP) increases a woman’s CV risk by 25%.^3^

There is now robust epidemiological evidence that pregnancy complications, like pre-eclampsia (PE) and gestational hypertension (GH), are associated with elevated CV risk and the risk of hypertension specifically - termed chronic hypertension (CH) in obstetric literature.^3^ The bulk of evidence suggests that this association is based primarily on shared underlying CV risk factors, rather than a direct damaging effect of the hypertensive disorder of pregnancy.^4^

Pregnancy offers a unique opportunity in a woman’s life to identify CV risk and take a life course approach to health promotion. Professional bodies now recommend that women with a history of PE or GH receive postpartum counselling about lifestyle and other CV risk factor modification, as well as long-term BP monitoring to detect CH.^5^ However, as clinical interventions are introduced that reduce the incidence of PE or GH, using these clinical outcomes alone to ‘flag’ women for long-term follow-up will become increasingly ineffective. For example, first trimester screening and aspirin therapy for women at increased risk of preterm PE (with delivery before 37 weeks’ gestation) reduces its incidence by more than 60%.^6^ Similarly, screening for PE risk at 35-36 weeks’ gestation and risk-stratified timing of birth may reduce the incidence of term PE by almost 60%.^7^ Additionally, for all pregnant women regardless of PE risk, the incidence of PE and GH is reduced by 40% by exercise,^8^ 50% by calcium,^9^ and 35% by timed birth at 39 weeks (for nulliparous women).^10^

In this study, we aimed to develop a predictive model for development of future CH, based on information collected in an index pregnancy.

## METHODS

### Study population

This was a prospective cohort study of women who attended routine early pregnancy hospital visits for PE screening at King’s College Hospital, London and Medway Maritime Hospital, Gillingham, UK, in two consecutive pregnancies, between March 2006 and August 2021.

Inclusion criteria were singleton pregnancies and delivery of a non-malformed liveborn or stillborn at ≥24 weeks in the index pregnancy. We excluded pregnancies with CH in the index pregnancy, and pregnancies ending in miscarriage, termination before 24 weeks of gestation, or those with known aneuploidies or major fetal abnormalities. All women gave written informed consent to participate in the screening study, which was approved by the NHS Research Ethics Committee (02-03-033). The data that support the findings of this study are available from the corresponding author upon reasonable request and pending Departmental approval.

In the index pregnancy, the early pregnancy visit was at 11+0 to 13+6 weeks’ gestation, and included: recording of maternal demographics (including self-identified ethnicity) and medical history; measurement of weight and height; and measurement of systolic and diastolic BP by validated automated devices and according to a standardised protocol.^11^ Gestational age was determined by measurement of fetal crown-rump length at 11-13 weeks’ gestation or the fetal head circumference at 19-24 weeks.

Outcome data were collected from hospital maternity or general medical practitioners’ records. PE was defined as per the American College of Obstetricians and Gynecologists,^12^ as CH or GH, and development of at least one of: new-onset proteinuria, serum creatinine >97 µmol/L in the absence of underlying renal disease, serum transaminases more than twice normal (≥65 IU/L for our laboratory), platelet count <100,000 / µL, headache or visual symptoms, or pulmonary oedema. CH was systolic BP ≥140mmHg or diastolic BP ≥90mmHg, at least twice, four hours apart or on two consecutive outpatient visits, and documented before pregnancy or 20 weeks’ gestation; the diagnosis was confirmed at the second trimester routine ultrasound scan for fetal anomalies. GH was defined as new-onset hypertension developing at ≥20 weeks’ gestation in a previously normotensive woman.^12^

### Outcome measure

The outcome for predictive modelling was ‘future’ CH, defined as above and in the subsequent pregnancy^13^. The diagnosis was confirmed at 20 weeks’ gestation, at the routine anatomy ultrasound scan, based on review of all BP measurements in pregnancy to that point.

### Statistical analysis

Data were summarised descriptively for women in the index pregnancy, according to whether they developed future CH (subsequent pregnancy). Median and interquartile range (IQR) were used for continuous variables and number (percentage) for categorical variables. The groups were compared by independent sample t-test for continuous variables, and chi-square test for categorical variables.

Logistic regression models were fitted, with identification of future CH (subsequent pregnancy) as the outcome, and as covariates, index pregnancy maternal characteristics, medical history, early pregnancy BP, and development of PE or GH. Functional forms were selected by plotting the proportion of future CH against grouped continuous covariates. Models were selected using a stepwise algorithm, to minimise the Akaike information criterion^18^. Predictive performance of the model was assessed by area-under-the-receiver-operator-characteristic (ROC) curve (AUC), and detection rates (DRs) for fixed screen-positive rates (SPRs).

The calibration of the predicted probabilities from the model, or risks, was assessed:

i. visually, by plotting the observed proportion of CH against the predicted risk; and
ii. quantitively, by recording measurements of calibration-in-the-large and calibration slope. Calibration-in-the-large is a measure of whether in general, the predicted risks are too high or too low, quantified by the estimated intercept from a logistic regression of incidence on the logit of risk, with the slope fixed at 1. The intercept is a measure of the deviation of the observed incidence from the predicted. For perfectly calibrated risks, the intercept should be zero. The calibration slope assesses the calibration across the range of risks and is the slope of the regression line of the logistic regression of incidence on the logit of risk; if the risk is well-calibrated, the slope should be 1.0.Five-fold cross validation was further used to assess risk calibration and predictive performance. The data were divided into five equal sub-groups. The model was then fitted five times to different combinations of four of five subgroups and used to predict the risk of developing future CH in the remaining one-fifth of the data.

Finally, the covariates from the final model were broken down into three components, maternal characteristics and medical history, early pregnancy BP, and index pregnancy outcome, and. Predictive performance of the final model was assessed against combinations of these component models to identify the benefit of index pregnancy PE or GH on predictive performance. These models were also compared with the detection rate for future CH using only PE or GH from the index pregnancy. All statistical analyses were performed with R statistical software. The package pROC was used for the ROC curve analysis, and the package PropCIs to calculate confidence intervals (CIs).

## RESULTS

### Study participants

The study population consisted of 26,511 women without CH in their index pregnancy, who were seen again for care in the subsequent pregnancy. In the index pregnancy, 1560 (5.9%) women developed either PE (760, 2.9%) or GH (800, 3.0%). A median [IQR] of 3.0 [1.6-5.0] years later by the time of their subsequent pregnancy, 215 (0.8%) women had developed CH.

At 11-13 weeks’ gestation in the index pregnancy, women who developed future CH (vs. those who did not) were different with regards to most baseline characteristics (Table 1). They were older and heavier. They were more often: self-identified as being non-White, particularly Black and South Asian ethnicities; had chronic medical conditions associated with PE risk (e.g., pre-gestational diabetes mellitus and systemic lupus erythematosus/antiphospholipid antibody syndrome); and had a family history of PE. Also, women who developed future CH (vs. those who did not) were more often parous, and had higher systolic and diastolic BP, with just under 20% (vs. about 1%) having a non-sustained BP ≥140/90mmHg (recorded only at a single visit) and about 1/3 (vs. just under 10%) a non-sustained BP of 130-139/80-89 mmHg.

**Table 1.** Characteristics of the study population <u>during the INDEX pregnancy</u>, according to development of future CH

| Characteristics and outcome in INDEX pregnancy | Did not develop future CH (n=26,296) | Developed future CH (n=215) | p-value |
| --- | --- | --- | --- |
| Preeclampsia (PE) | 709 (2.7) | 51 (23.7) | <0.0001 |
| Gestational hypertension (GH) | 739 (2.8) | 61 (28.4) | <0.0001 |
| Unaffected by PE or GH | 24,848 (94.5) | 103 (47.9) | <0.0001 |
| Maternal age (years) | 28.9 (24.3, 32.5) | 30.4 (26.8, 34.25) | <0.0001 |
| Maternal weight (kg) | 65.1 (58.1, 75.0) | 80.0 (66.85, 93.5) | <0.0001 |
| Maternal height (cm) | 165 (160, 169) | 165 (160, 170) | 0.75 |
| Body mass index (kg/m<sup>2</sup>) | 24.0 (21.6, 27.6) | 29.4 (25.2, 33.7) | <0.0001 |
| Gestational age (days) | 88.9 (86.2, 91.7) | 88.7 (85.5, 91.7) | 0.30 |
| Ethnic origin |  |  | <0.0001 |
| White | 20,608 (78.4) | 91 (42.3) |  |
| Black | 3,585 (13.6) | 109 (50.7) |  |
| South Asian | 1,205 (4.6) | 10 (4.7) |  |
| East Asian | 352 (1.3) | 2 (0.9) |  |
| Mixed | 546 (2.1) | 3 (1.4) |  |
| Diabetes mellitus type 1 | 126 (0.5) | 2 (0.9) | 0.01 |
| Diabetes mellitus type 2 | 73 (0.3) | 3 (1.4) | 0.01 |
| SLE/APS | 95 (0.4) | 3 (1.4) | 0.05 |
| Smoker | 2,569 (9.8) | 14 (6.5) | 0.14 |
| Family history of PE | 1,821 (6.9) | 34 (15.8) | <0.0001 |
| Method of conception |  |  | 0.74 |
| Spontaneous | 25,373 (96.5) | 209 (97.2) |  |
| *In vitro* fertilisation | 569 (2.2) | 3 (1.4) |  |
| Ovulation drugs | 354 (1.4) | 3 (1.4) |  |
| Parity |  |  | <0.0001 |
| Parous, no previous PE | 5,221 (19.9) | 64 (29.8) |  |
| Parous, previous PE | 332 (1.3) | 20 (9.3) |  |
| Nulliparous | 20,743 (78.9) | 131 (60.9) |  |
| Interpregnancy interval (years) | 2.3 (1.4, 3.7) | 3.0 (1.6, 5.0) | 0.0002 |
| Mean arterial pressure | 85.3 (80.3, 90.8) | 98.5 (91.7, 102.9) | <0.0001 |
| Systolic blood pressure | 115.5 (108.8, 122.8) | 129.5 (122.15, 137.4) | <0.0001 |
| <110 mmHg | 10 (4.7) | 7779 (29.6) |  |
| [110-120) mmHg | 29 (13.5) | 9486 (36.1) |  |
| [120-130) mmHg | 73 (34.0) | 6677 (25.4) |  |
| [130-140) mmHg | 63 (29.3) | 2010 (7.6) |  |
| ≥140 mmHg † | 40 (18.6) | 344 (1.3)‡ |  |
| Diastolic blood pressure | 70.3 (65.3, 75.5) | 81.5 (74.65, 87.0) | <0.0001 |
| <70 mmHg | 12625 (48.0) | 25 (11.6) |  |
| [70-80) mmHg | 10672 (40.6) | 66 (30.7) |  |
| [80-90) mmHg | 2766 (10.5) | 86 (40.0) |  |
| ≥90 mmHg † | 233 (0.9) | 38 (17.7) |  |

APS = antiphospholipid syndrome; BP = blood pressure; CH = chronic hypertension; GH = gestational hypertension; MAP = mean arterial pressure; MoM = multiples of the median; PAPP-A = pregnancy-associated plasma protein-A; PE = pre-eclampsia; PIGF = placental growth factor; SLE = systemic lupus erythematosus; Ut-PI = uterine artery pulsatility index.
† Women with systolic BP ≥140mmHg or diastolic BP ≥90mmHg at the 11-13 weeks' gestation screening visit, were followed in the index pregnancy, and none had BP sustained at this level that would be consistent with CH.

Women who developed future CH (vs. those who did not) more often developed PE or GH in their index pregnancy (Table 1).

### Predictors of development of CH

The best-fitting model for prediction of future CH (Table 2) included terms for: maternal age, weight, systolic and diastolic BP, parity, family history of PE, GH and the gestational age at delivery with PE.

**Table 2.** Fitted model using characteristics and outcome in the INDEX pregnancy for prediction of future CH

| Characteristics and outcomes in INDEX pregnancy | Odds ratio (95% CI) | Estimate |
| --- | --- | --- |
| Intercept |  | -7.6239 |
| PE | 18.8 (6.0 -58.9) | 2.93 |
| Gestational age at delivery with PE in weeks - 24 | 0.9306 (0.8578 - 1.0096) | -0.07 |
| GH | 8.1893 (5.6608 -11.8471) | 2.10 |
| Maternal weight in kg - 60 | 1.0165 (1.0084 - 1.0246) | 0.02 |
| Maternal age in years - 30 | 1.0384 (1.0090 - 1.0685) | 0.04 |
| Black ethnicity | 5.1841 (3.7732 - 7.1224) | 1.66 |
| South Asian ethnicity | 2.9688 (1.4865 - 5.9295) | 1.09 |
| Family history of PE | 1.7982 (1.1894 - 2.7185) | 0.59 |
| Parous, no previous PE | 1.6946 (1.1906- 2.4122) | 0.53 |
| Parous, previous PE | 2.7851 (1.5572- 4.9811) | 1.02 |
| sBP in mmHg - 110 if sBP >110; 0 if sBP ≤110 | 1.1015 (1.0575- 1.1474) | 0.10 |
| (sBP in mmHg - 110)^2 if sBP >110; 0 if sBP ≤110 | 0.9990 (0.9981- 0.9999) | -0.001 |
| dBp in mmHg - 75 if dBp >75; 0 if dBp ≤75 | 1.0979 (1.0655- 1.1313) | 0.09 |

BP = blood pressure; CH = chronic hypertension; dBp = diastolic BP; GH = gestational hypertension; PE = pre-eclampsia; sBP = systolic BP

Figure 1 shows the observed proportions of future CH for continuous variables, presented with fitted lines by grouped maternal age, weight and BP; ‘broken stick’ relationships were fitted for both systolic and diastolic BP, with the breaks at 110 and 75 mmHg, respectively.

**Figure 1.**
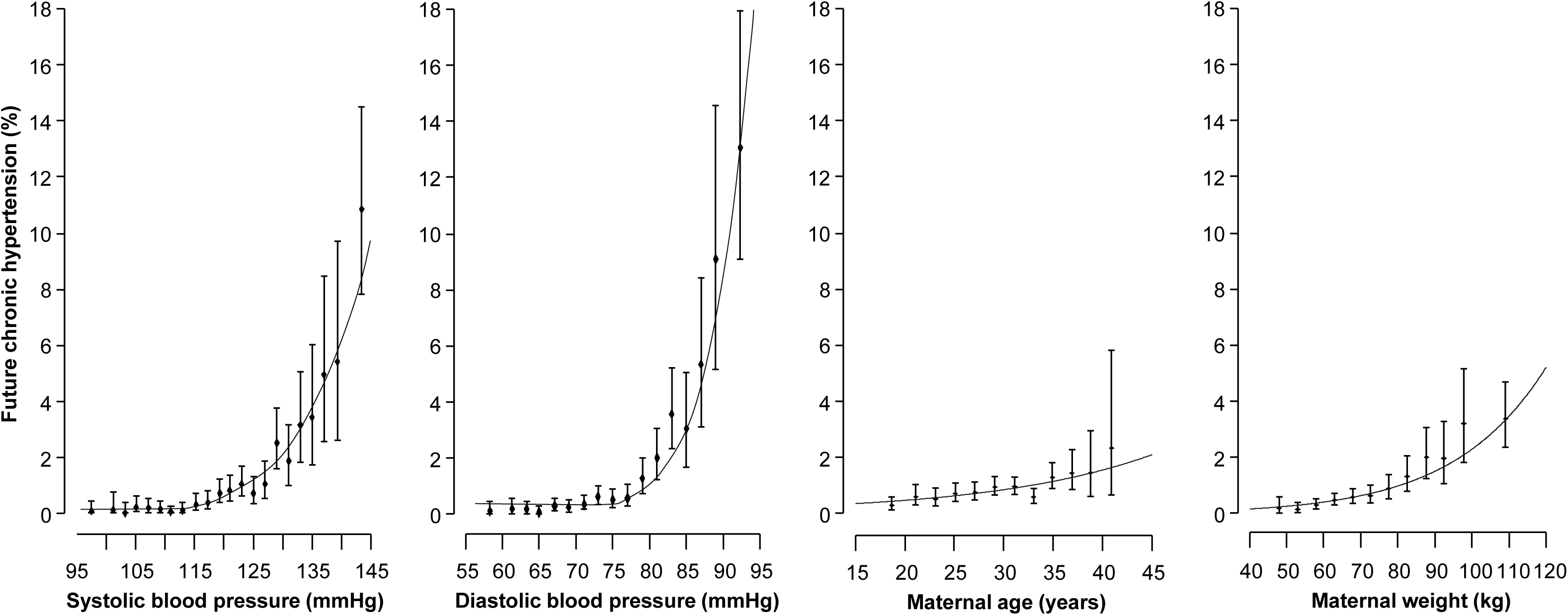
Relationship between first trimester systolic and diastolic blood pressure, maternal age and weight with observed proportions of future chronic hypertension.

Figure 2 presents the odds ratios (ORs) with 95% CIs for development of future CH for categorical variables (ethnicity, parity, family history of PE, and development of PE or GH in the index pregnancy), ordered by their strength of association with future CH. Of note, while the risk of future CH was higher for parous (vs. nulliparous) women, the risk was particularly high for parous women with prior PE (vs. parous women with no such history). The strongest risk factors for future CH were Black ethnicity, GH, and PE. While future CH risk was similar after PE or GH, the risk was higher the earlier the woman delivered with PE. The five-fold increased risk of future CH associated with delivery with PE at 42 weeks was similar in magnitude to the risk for women of Black ethnicity.

**Figure 2.**
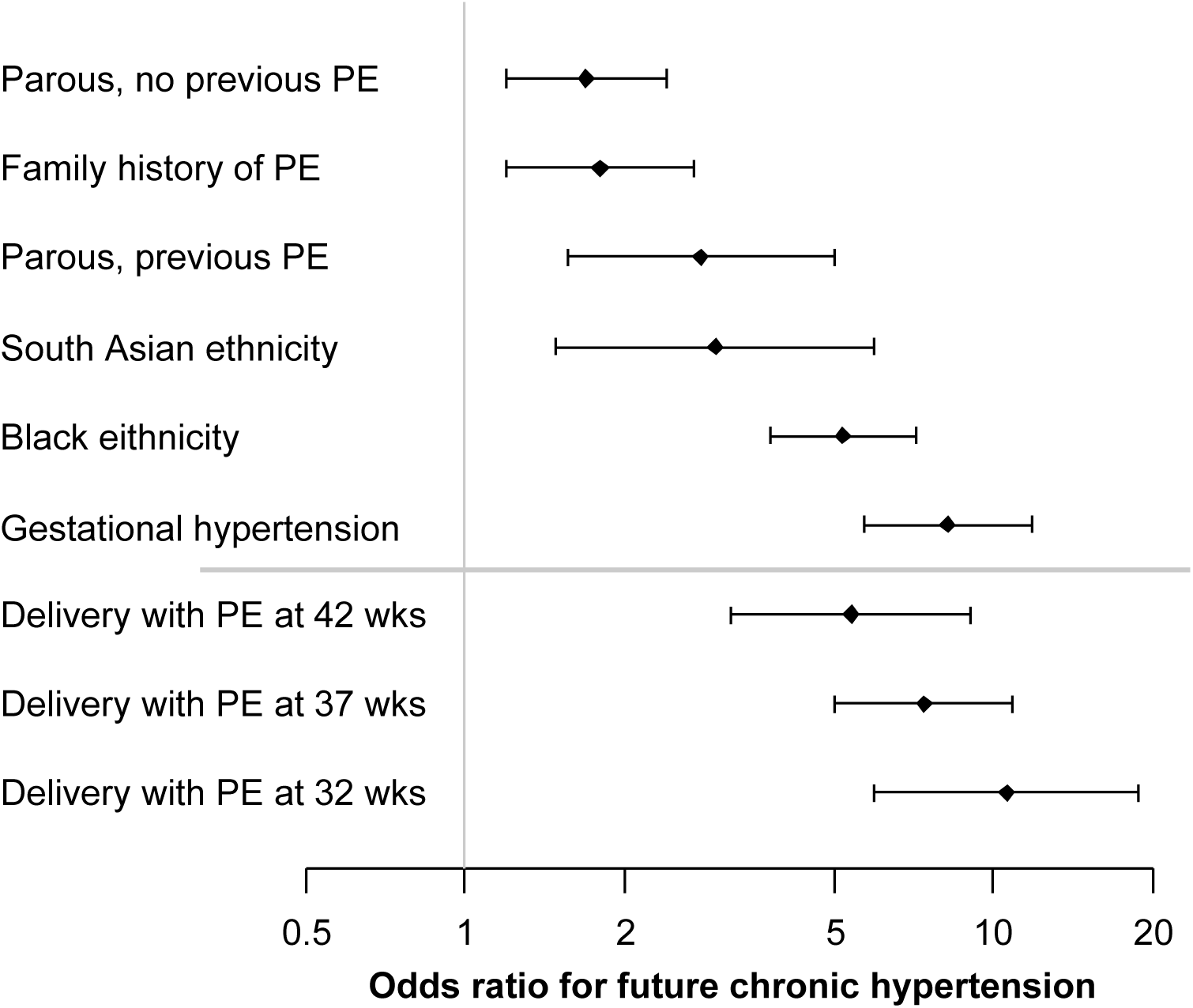
Forest plot of maternal and pregnancy characteristics with odds for development of future chronic hypertension. PE – pre-eclampsia

### Predictive performance and calibration

The 1560 (5.9%) women who developed PE or GH in the index pregnancy, accounted for 112 cases of future CH, for a detection rate of 52.1% (Table 3). For the same SPR of 5.9%, the model for prediction of future CH had a similar detection rate of 54.0% when based on maternal characteristics and early pregnancy BP, but the detection rate (67.0%) was improved when development of PE or GH was added to the model (Table 3).

**Table 3.** Performance of screening in pregnancy for prediction of future CH

| Method of screening | N women;<br>DR at 5.9% SPR | N women;<br>DR at 10% SPR | AUROC<br>(95% CI) |
| --- | --- | --- | --- |
| PE or GH alone | 112; `52.1 (45.2-58.9) | Not applicable | Not applicable |
| Multivariable model |  |  |  |
| Maternal characteristics | 84; 39.1 (32.5-45.9) | 103; 47.9 (41.1 - 54.8) | 0.81 (0.77-0.84) |
| + BP in early pregnancy | 116; 54.0 (47.0-60.8) | 146; 67.9 (61.2 - 74.1) | 0.90 (0.88-0.92) |
| + PE or GH | 128; 59.5 (52.6-66.2) | 147; 68.4 (61.7 - 74.5) | 0.88 (0.86-0.91) |
| + BP in early pregnancy<br>+ PE or GH | 144; 67.0 (60.3-73.2) | 158; 73.5 (67.1 - 79.3) | 0.93 (0.91-0.94)* |

AUROC = area-under-the-receiver-operator characteristic; BP = blood pressure; CH = chronic hypertension; DR = detection rate; GH = gestational hypertension; N = number of new cases of CH detected from a total of 215; PE = pre-eclampsia; SPR = screen-positive rate.
\* PE risk was determined by the multivariable competing risks model at 11-13 weeks' gestation, using placental growth factor or if unavailable, pregnancy-associated plasma protein-A (https://fetalmedicine.org/research/assess/preeclampsia/).
† Significantly better than screening by maternal characteristics plus either BP in early pregnancy or 'PE or GH'.

**Table 4.** Five-fold cross validation. Area-under-the-curve, intercept and slope with 95% confidence intervals

| **Fold** | **Area-under-the-curve** | **Intercept** | **Slope** |
| --- | --- | --- | --- |
| 1 | 0.9357 (0.9009-0.9705) | -0.0600 (-0.3920-0.2721) | 1.0358 (0.8629-1.2088) |
| 2 | 0.9345 (0.9029-0.9661) | 0.0397 (-0.2881-0.3675) | 1.0832 (0.8944-1.2720) |
| 3 | 0.9072 (0.8726-0.9419) | 0.1708 (-0.1353-0.4770) | 0.8549 (0.7116-0.9982) |
| 4 | 0.9042 (0.8468-0.9616) | 0.0103 (-0.3173-0.3379) | 0.9794 (0.8126-1.1462) |
| 5 | 0.9165 (0.8761-0.9568) | -0.1762 (-0.5139-0.1615) | 0.8980 (0.7434-1.0526) |

For a SPR of 10%, the model for prediction of future CH performed similarly when including maternal characteristics and either early pregnancy BP (detection rate of 67.9%) or development of PE or GH (68.4%) (Table 3). Inclusion of all predictors (i.e., maternal characteristics, early pregnancy BP, and development of PE or GH) optimised detection of future CH (detection rate 73.5%) and had the best model performance in terms of AUC (0.93, 95% CI 0.91-0.94). ROC curves are presented graphically in Figure 3.

**Figure 3.**
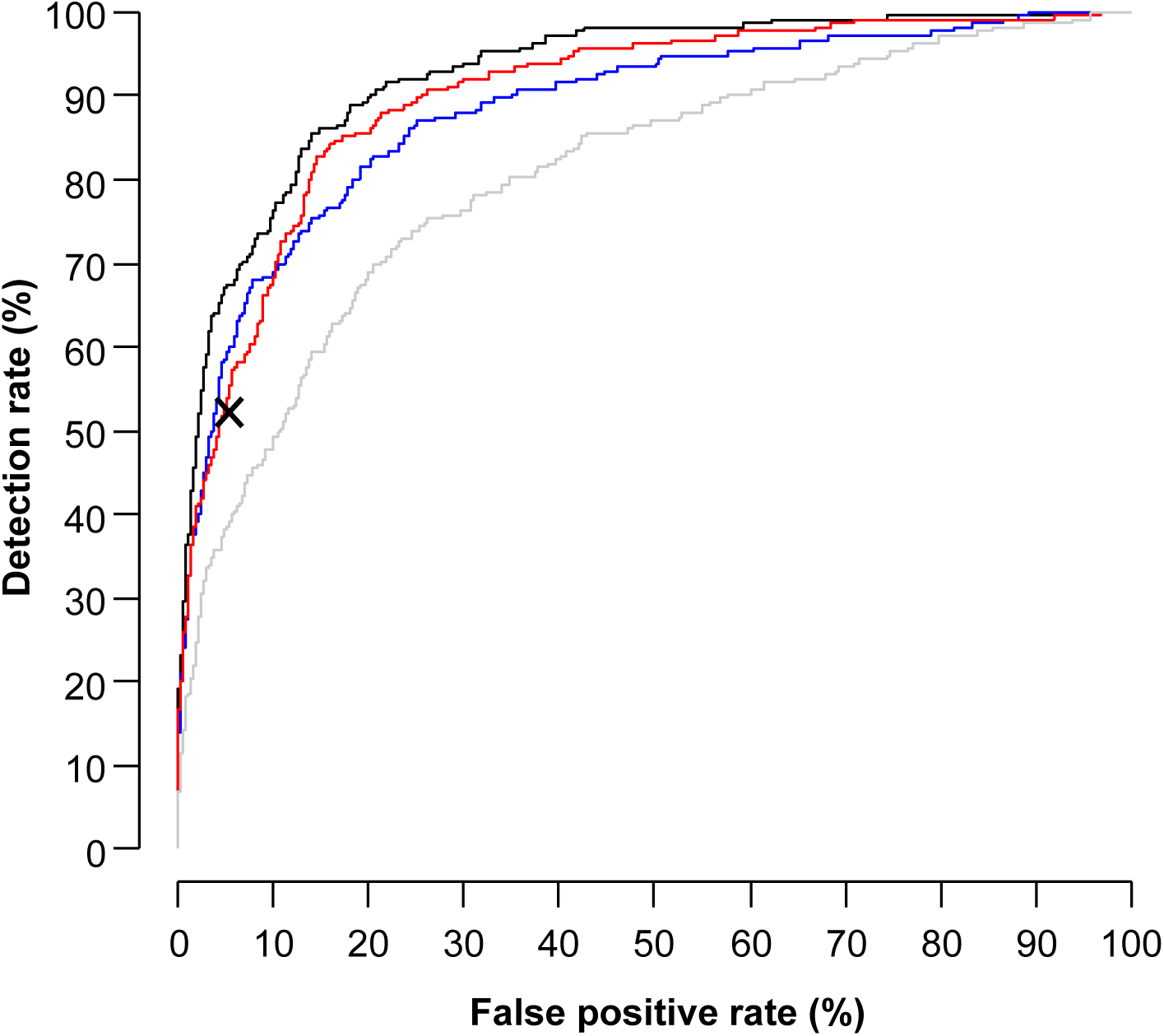
Receiver-operating-characteristic curves for screening for future chronic hypertension by maternal characteristics (grey curve), combination of maternal characteristics and preeclampsia (PE) or gestational hypertension (GH) in the index pregnancy (blue curve), maternal characteristics and early pregnancy blood pressure (red curve), or a combination of maternal characteristics, early pregnancy blood pressure, and PE or GH in the index pregnancy (black curve). The ‘X’ represents the detection rate and false positive rate of screening by PE or GH alone.

**Figure 4.**
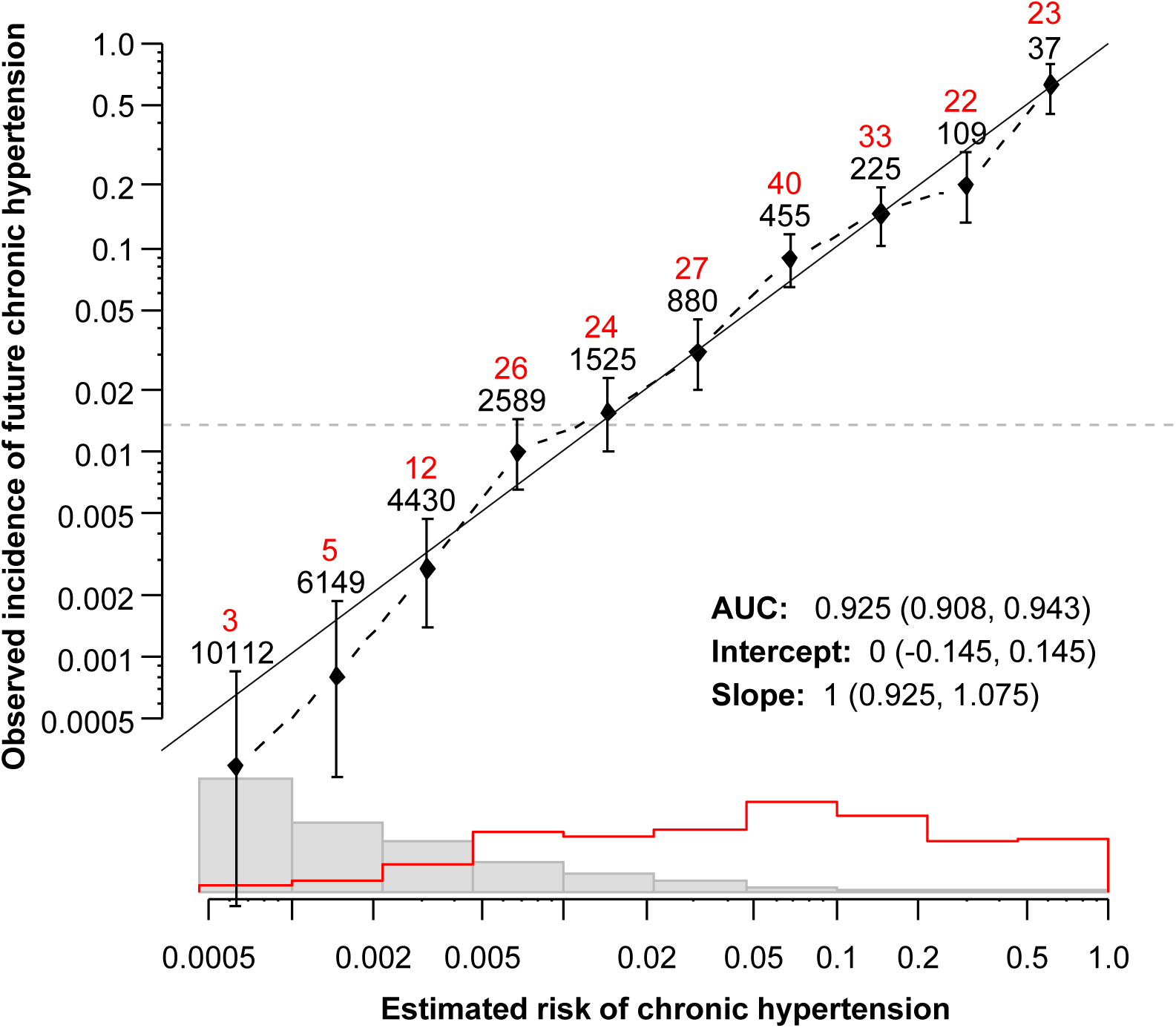
Calibration plots for the best-performing model for prediction of future chronic hypertension. The diagonal black line is the line of perfect agreement. The overall incidence of future chronic hypertension is shown by the horizontal interrupted line. The diamonds and vertical lines are the observed incidence of future chronic hypertension with 95% confidence intervals, the numbers in red are the observed number of cases of future chronic hypertension, and the numbers in black are the number of cases in each estimated risk category. The histograms show the distribution of risks in index pregnancies after which future chronic hypertension develops (red), compared with those after which no chronic hypertension develops (grey).

For the full data set, the calibration intercept was 0 and the slope 1. Results from the five-fold cross validation are consistent with this, with intercepts being close to 0 and slopes close to 1 for all 5 folds.

## DISCUSSION

### Main findings

In this prospective, non-intervention screening study of 26,511 women from socioeconomically and ethnically diverse South East England, we have shown that early pregnancy characteristics can identify women at increased risk of future CH, defined in this study during their subsequent pregnancy, on average three years later.

Our multivariable model of maternal characteristics (age, weight, ethnicity, parity, and family history of PE) and early pregnancy BP (at 11-13 weeks’ gestation) performed similarly to knowledge of the development of PE and GH later in the same pregnancy. As early pregnancy characteristics will be unaffected by interventions to reduce PE and GH, our data provide a useful model to identify women at risk for future CH who may benefit from postpartum follow-up, as currently recommended only for women with prior PE or GH. The model provided a very high discrimination between affected and unaffected pregnancies and there was good agreement between the predicted risks and observed incidence of future CH.

### Interpretation in light of existing evidence

The risk factors for future CH that we identified in early pregnancy fit with those identified outside pregnancy. In our study population, non-sustained BP in the 130-139/80-89 mmHg range in the index pregnancy was common, and a strong risk factor for development of future CH. From our analysis, it was also apparent that the risk for future CH increased exponentially as women’s BP approached a prehypertensive state. A continuous relationship between BP and development of hypertension is well-documented outside pregnancy, and BP is known to track across the life course. For example, women often experience a steep rise in BP during the third decade of life, with this trend continuing thereafter,^14^ and systolic BP in early adulthood is associated with future CV risk.^15^ Also, women of Black (vs. White) ethnicity have a higher BP phenotype outside pregnancy;^16^ the mechanisms responsible remain poorly defined, and are often attributed to differences in genetics, socioeconomic status, environmental conditions, and/or barriers to healthcare delivery and access.^16^

The association in our study between parity (even without a history of PE) and a higher risk of future CH may seem counterintuitive. However, this finding is consistent with a large meta-analysis (10 cohort studies, over 3 million women) that demonstrated an association between parity (vs. nulliparity) and more CV events.^17^ The reasons for greater CV risk associated with parity are uncertain; potential biologic mechanisms include unfavourable pregnancy-related changes in CV risk factors (such as lipids, glucose, and weight), as well as endothelial dysfunction and inflammatory and haemostatic processes.

Our findings confirm an association between prior PE or GH, and the risk of future CH, including similar odds after PE or GH (7-8 times higher compared with previously normotensive pregnancy). Similar results were seen after three years’ mean follow-up in the recent French nationwide CONCEPTION study involving 2,663,573 pregnancies.^18^ In other published studies, the magnitude of future CH risk following PE or GH has varied, but is evident within a few years of birth: 12-15 times higher in the first year or 4-10 times higher at one to five years (Danish register-based cohort, 1.5M women), 2.7 times higher within three years (observational study, 4484 women); and 2.8 times higher within five years (Nurses’ Health Study II, 58,671 women).^19^ In a meta-analysis of 25 studies and 3 million women, women who developed PE (vs. those with a normotensive pregnancy), had a 3.7 times higher risk of future CH over a period of 14 years.^17^ This variation in CH incidence may be related to between-study differences in risk factors (such as ethnicity), and/or the duration of study follow-up. Regardless, the fact that both PE and GH are related to CH risk is consistent with their associations with elevated CV risk (umbrella review of 2 systematic reviews including 4 cohort studies^20^.

Timing of PE development is a strong risk factor for development of cardiac disease and CH. Our findings were similar, in that women who delivered with PE at 32 weeks had almost double the risk for future CH, compared with those who delivered with PE at 42 weeks. Differences in the pathophysiology of term vs preterm PE have not been clearly defined, however, it is well-known that preterm (vs. term) PE occurs less frequently and has a more severe phenotype, with placental insufficiency and increased risk for maternal morbidity and mortality.^21^

Our findings are consistent with epidemiological studies suggesting that shared underlying risk factors drive the association between PE or GH and the risk of future CH and CV disease. These shared risk factors may be known health conditions (such as obesity), or pre-existing but subclinical abnormalities in cardiac and vascular health (such as increased peripheral vascular resistance or endothelial dysfunction). It is theoretically possible that PE or GH may themselves damage the maternal CV system, but there is little evidence to support this concern First, adjusting for conventional CV risk factors eliminates entirely or almost entirely any association observed between PE or GH and CV disease, although this was not the case in a recent Swedish registry study examining coronary atherosclerosis by computed tomograph^22^ Second, prolonging maternal exposure to PE through its expectant care has yielded inconsistent estimates of the effect on CV disease.^23^ Third, adding a history of PE or GH to traditional 10-year CV risk estimate algorithms has changed their performance little or not at all^24^

### Implications for clinical practice

Most pregnancy hypertension clinical practice guidelines already recognise the link between PE or GH and elevated CV risk, including development of future CH. These guidelines recommend annual CV assessment, lifestyle counselling, and modification of CV risk factors postpartum. However, a key challenge that we face in moving forward, is that progress being made in the prevention of PE and GH will for many women, remove the ‘flag’ for postnatal follow-up; yet, the CV risk of women who no longer develop PE or GH will remain in the long-term.

Knowing that a woman is at heightened risk of future CH is of value, particularly when that risk can be identified in pregnancy, when women routinely seek health care. Many CV risk factors are modifiable and effective interventions from outside pregnancy include diet and lifestyle change, weight loss, smoking cessation, and a reduction in alcohol intake. Of additional and particular interest postpartum are prevention of pregnancy weight retention,^25^ promotion of breastfeeding for maternal health,^26^ and potentially, BP self-management.^27^ In a pilot randomised trial (91 women), BP self-management consistent with UK national guidance to maintain BP <150/100mmHg (vs. usual care), resulted in significantly lower BP, well after medications had been stopped; the reduction in BP documented at six months postpartum was confirmed to be sustained for another three years (61 women)^28^. If a short period of BP control were confirmed to result in sustained lowering of BP and improvement in a woman’s BP trajectory, this would represent a highly-attractive strategy to address CV risk.

### Strengths and limitations

Our study’s strengths include use of data from a well-characterised, ethnically diverse cohort with detailed reporting of maternal and pregnancy clinical and demographic characteristics, BP measurement by trained physicians using a well-established protocol, and diagnosis of PE, GH, and future CH according to accepted definitions.^12, 13^ We used information that is regularly obtained as part of first trimester screening for PE in Fetal Medicine Units. The diagnosis of CH was based not only on use of antihypertensive medication or single elevated BP values early in the subsequent pregnancy, but on serial and protocolised measurement of BP at <20 weeks’ gestation (as above).

Limitations of our study include our lack of interpregnancy BP measurements, and therefore, the timing of onset of future CH that developed by the subsequent pregnancy. Also, our model for prediction of future CH was derived from our population and may not be applicable to other populations with different characteristics, such as less ethnic diversity.

### Conclusions

To capitalise on health promotion opportunities in pregnancy, and identify women at increased risk of future CH, we suggest targeting early pregnancy maternal characteristics and BP. Not only do these variables perform similarly to development of PE or GH (later in the index pregnancy) in identifying women at future CH risk, but they will be unaffected by interventions that reduce PE or GH and remain a stable target for CV risk reduction interventions over time.

**Acknowledgments:** The ultrasound machines for fetal echocardiography and the software for speckle tracking analysis were provided free-of-charge by Canon Medical Systems Europe BV, Zoetermeer, The Netherlands. These bodies had no involvement in the study design; in the collection, analysis and interpretation of data; in the writing of the report; and in the decision to submit the article for publication.

**Funding:** The study was supported by a grant from the Fetal Medicine Foundation (Charity No: 1037116).

### **Declaration of interests**: The authors had no interests to declare

## Data Availability

The data can be made available upon request

